# Penetration of Inner Scleral Fibers into Peripapillary Sclera Revealed by Polarization-Sensitive OCT

**DOI:** 10.1101/2025.02.22.25322543

**Authors:** Jianping Xiong, Hongshuang Lu, Nan Zhou, Yijin Wu, Masahiro Yamanari, Hiroyuki Takahashi, Michiaki Okamoto, Keigo Sugisawa, Takeshi Yoshida, Changyu Chen, Yining Wang, Ziye Wang, Kyoko Ohno-Matsui

## Abstract

**Purpose:** To determine the structural characteristics of the peripapillary sclera in myopic eyes using polarization-sensitive optical coherence tomography (PS-OCT).

**Methods:** Patients with myopia who underwent PS-OCT imaging between April and June 2023 were studied. The intensity, optic axis, and birefringence of PS-OCT images were analyzed to determine the surface of the lamina cribrosa (LC). The direction and arrangement of the scleral collagen fibers in the peripapillary region and scleral ridge were determined.

**Results:** One hundred and ten eyes of 59 patients with myopia (mean axial length, 30.00 ± 1.98 mm) were studied. The scleral fibers around the optic nerve were arranged circularly in all eyes regardless of the presence or size of the peripapillary atrophy. Inner scleral fibers that penetrated beneath this circular arrangement were detected in 98 eyes (89.1%). There was a Y-shaped split at the point of penetration in 77 eyes (70.0%). The integrity of the circular arrangement and penetrating scleral fiber complexes remained intact even in cases of severe peripapillary scleral deformation. A scleral ridge was identified in 27 eyes (24.5%), and it appeared as a full-thickness protrusion composed primarily of outer scleral fibers. The ridge was consistently located outside the circular arrangement.

**Conclusions:** The penetration of the radial inner scleral fibers into the circular arrangement and extension to the LC suggest that they act as stabilizing anchors for the circular structure. The differences of the scleral ridges from those in eyes with dome-shaped macula suggest that unique mechanisms cause the scleral differences.

## Introduction

The peripapillary sclera plays a critical role in the biomechanics of the eye.^1–3^ Its structure (shape and thickness) and material properties (stiffness, brittleness, and compliance) affect the mechanical sturdiness of the optic nerve (ON) significantly. These properties thereby affect its susceptibility to increased intraocular pressure (IOP) and other internal and external pressures.^4^

Earlier studies examined the structure of the peripapillary sclera by wide-angle X-ray scattering (WAXS), small-angle light scattering (SALS), and second harmonic generation microscopic images of normal human and rat eyes.^5,6^ These studies identified a key feature of the posterior scleral structure: a ring of collagen fibers encircling the scleral canal. Gogola et al. used polarized light microscopy (PLM) to show a radial arrangement of the inner scleral fibers and circumferential arrangement of the outer scleral fibers. These findings were consistent in humans, monkeys, pigs, cows, goats, and sheep^7^, and they have greatly enhanced our understanding of the anatomy of the peripapillary sclera.

Polarization-sensitive optical coherence tomography (PS-OCT) is an advanced OCT technology that uses polarized light as an additional image contrast mechanism.^8,9^ It uses the birefringent properties of the sclera to image and quantify the variations of birefringence across different regions of the sclera. PS-OCT can also make visible the axis of orientation, or optic axis, of the birefringence associated with the direction of the fiber bundles.^10^ Based on these capabilities of PS-OCT, Baumann et al.^11^ in rats and Willemse et al.^12^ in healthy humans eyes to confirm the peripapillary scleral 3D structure. Most earlier studies suggested that the radial inner scleral fibers terminated at the circular arrangement adjacent to the scleral canal.^5–7,11–14^

The peripapillary circular scleral structure is known to provide protection and support to the ON.^5,7,13,14^ Thus, Hua et al. created seven posterior pole models demonstrating that radial fibers also contribute to the support of the posterior ocular surface.^1^ Most importantly, they demonstrated that the combination of the radial and circumferential fibers provided enhanced ON protection at elevated IOP. However, the structural relationship between the peripapillary scleral fibers of different orientations has not been well explored using *in vivo* imaging. This deficiency is due to the difficulties in obtaining detailed images of the peripapillary scleral structures *in vivo*. However, the choroidal thinning in myopic eyes facilitates the ability to record clear images of all the scleral layers by PS-OCT.

Thus, the purpose of this study was to determine the structural construction of the peripapillary sclera in myopic human eyes. To accomplish this, we analyzed the structure of the sclera in the PS-OCT images of myopic eyes.

## Methods

The procedures used in this study conformed to the tenets of the Declaration of Helsinki and were approved by the Ethics Committee of the Institute of Science Tokyo (Science Tokyo). The procedures complied with the standards for case series studies which allowed us to present the results thoroughly and transparently.

We reviewed the medical records of highly myopic patients who had undergone PS-OCT examinations between April 1, 2023, and June 16, 2023 at the Advanced Clinical Center for Myopia at Science Tokyo. Cases with poor-quality PS-OCT images were excluded. All patients had undergone a comprehensive ophthalmological examination including measurements of the axial length (IOL Master; Carl Zeiss Meditec AG, Jena, Germany), best-corrected visual acuity (BCVA), refractive error (spherical equivalent), color fundus photography, and swept-source OCT (SS-OCT, Triton; Topcon, Tokyo, Japan).

### PS-OCT Examinations and Image Processing

To assess the characteristics of the birefringence of the posterior segment of the eye, we developed a PS-OCT system using a swept-source laser centered at 1050 nm (ROCTIA, Tomey Corporation, Japan). The hardware of this system is commercially available in Japan. A comprehensive description of the technical concepts and the system has been presented.^15,16^ Retinal images were obtained over scan areas of 81 mm^2^ (9 × 9 mm) or 144 mm^2^ (12 × 12 mm) using a raster scanning protocol with 1024 A-scans along the horizontal axis and 256 B-scans along the vertical axis. In addition, a 6-line radial scan protocol was also used. This system has an axial resolution of 7.3 μm with an axial measurement range of 4.49 mm.

Already developed algorithms^10,17–19^ were adapted to process the measured raster scan data that yielded the polarization-diverse OCT intensities^20,21^, local phase retardation or magnitude of birefringence, and the orientation of the optic axis (OA) of the birefringence. These examinations resulted in three corresponding images: referred to as the intensity, birefringence, and OA images. To further enhance the image quality, the axial pixel separation was increased which improved the performance in localization of the axially accumulated Jones matrix.^10^ A necessary correction was used to resolve the OA at each axial depth.^10^

The birefringence of collagen fibers occurs when light passes obliquely or perpendicularly to the OA as in fundus imaging. This causes the collagen fibers to exhibit two refractive indices that alters the polarization state along the axial depth. In perfectly aligned fibers, the PS-OCT can mathematically determine the orientation of the OA on a plane perpendicular to the direction of the light.^10^ However, scleral collagen fibers have an interwoven structure that exceeds the resolution of OCT,^22^ and this results in a net birefringence from partially canceled birefringence of the individual fibers. PS-OCT provides depth-resolved measurements of this net birefringence *in vivo*. The OA, which corresponds to the preferential orientation of the net birefringence created by interwoven collagen fibers, reflects the preferential orientation of the fibers despite their complex interwoven structure.

For the data analysis, we processed the horizontal raster scan data and obtained three types of images with 256 cross-sections for each eye. The intensity images were used to locate the anterior surface of the lamina cribrosa (LC), the OA images were used to determine the direction of the scleral collagen fibers especially in the peripapillary area and at the scleral ridge. A scleral ridge was defined as an anterior protrusion of the sclera that is located temporal to the ON with a height greater than 150 μm.^23,24^ Radial scan images centered on the ON were used to determine the presence and height of the ridge. More specifically, the ridge measurements were made along the horizontal meridian using built-in tools (ROCTIA, Tomey Corporation, Japan). Birefringence images were used to assist in determining the alignment of the scleral fibers. High birefringence corresponded to more aligned scleral fibers^25^, and low birefringence corresponded to less aligned scleral fibers such as interwoven fibers.

Streamline rendering is a method for viewing the vectorial data and effectively makes visible the orientation of fibrous tissues such as the OA. This technique has been applied to imaging the OA of the heart muscles, articular cartilage, and scleral collagen fibers using PS-OCT.^10,18,26^ We used the ParaView software (version 5.12.0, Kitware Inc.) for volumetric streamline rendering of the OA following the methods of earlier research.^10^ The streamline images obtained directly and showed clearly the course of the scleral fibers.

### Statistical Analyses

Statistical analyses were performed using SPSS version 25.0 (IBM Corp., 2017. IBM SPSS Statistics for Windows, Version 25.0. Armonk, NY: IBM Corp.). Normally distributed data are expressed as the means ± standard deviations (SD), and non-normally distributed data are presented as medians with corresponding quartiles.

## Results

One hundred and ten eyes of 59 patients with myopia that had undergone PS-OCT imaging at the Advanced Clinical Center for Myopia at Science Tokyo between April 1, to June 16, 2023, were studied. There were 35 women and 24 men with a mean (± SD) age of 55.1 ± 10.2 years and range of 26.8 to 76.3 years. The mean axial length was 30.00 ± 1.98 mm with a range of 24.63 to 34.13 mm. The mean refractive error (spherical equivalent) was -13.24 ± 4.62 diopters (D) with a range of -1.63 to -24.75 D excluding 30 eyes with an implanted intraocular lens (IOL) and 1 eye with a contact lens. The median best-corrected visual acuity (BCVA) was 0.0 logMAR units with a range -0.3 to 1.0 logMAR units.

The *en face* OA images were revised by a one-dimensional moving average for the 15 axial pixels (67 µm) in the scleral slab, and the resulting images showed that the fibers in the inner layer of the sclera were distributed radially (Figure 1). The inner and outer layers of the sclera could be clearly distinguished in the horizontal cross-sectional OA images. The collagen fibers of the inner and outer layers of the sclera surrounding the optic nerve (ON) had different orientations: the inner scleral fibers extended radially from the ON to the periphery, while the outer scleral fibers were oriented vertically and obliquely (Figure S1).

**Figure 1.**
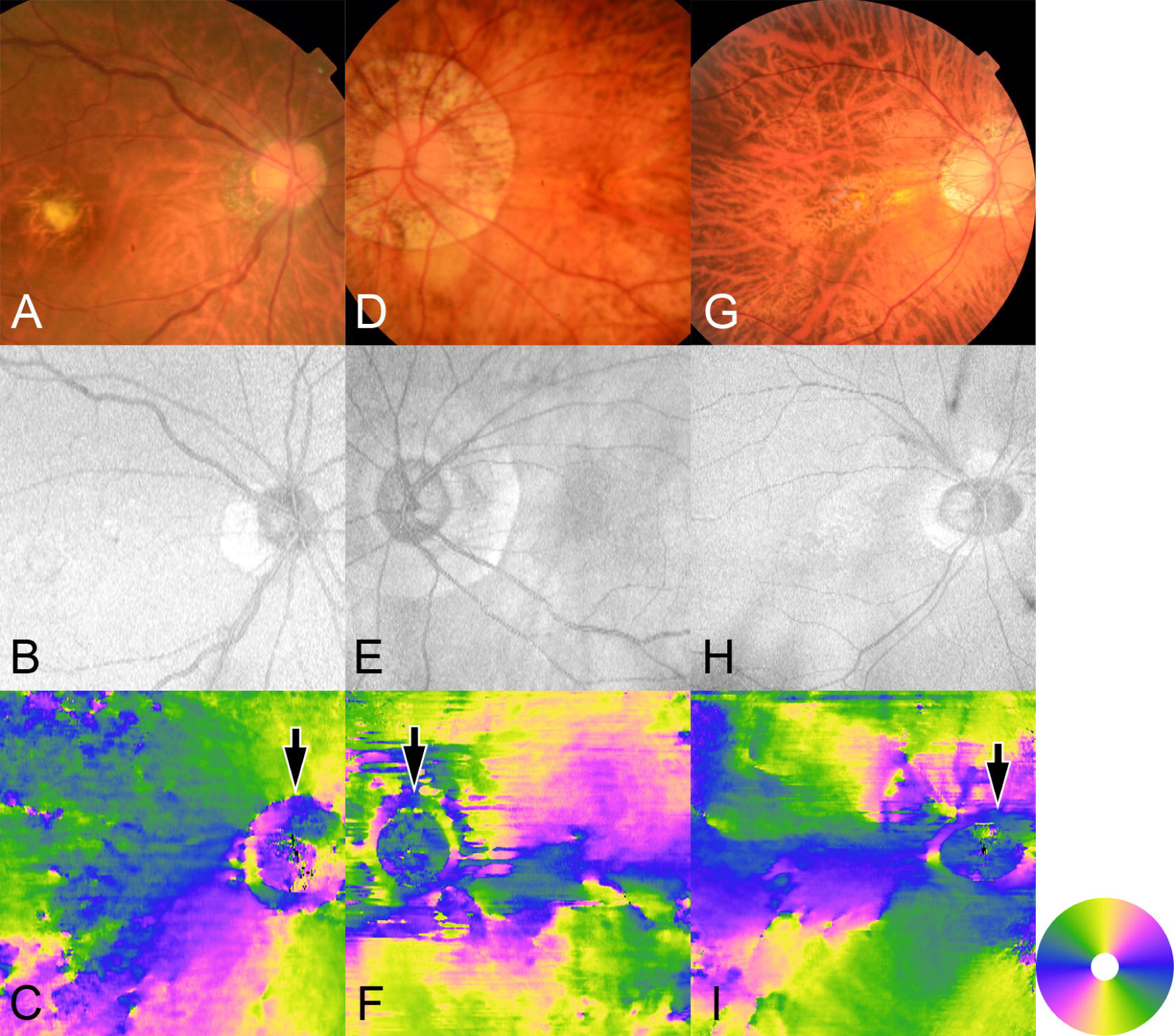
Circular arranged scleral fibers around the optic nerve (ON), and their relationship with the ON size rather than the peripapillary atrophic (PPA) size. A: Fundus photograph of the right eye of a 71-75-year-old woman with a refractive error (spherical equivalent) of -7.6 diopters (D) and an axial length (AL) of 25.7 mm with a tessellated fundus. Scarred macular neovascularization can be seen. B: *En face* optical coherence tomographic (OCT) intensity projection image shows a small PPA temporal to the ON. C: The optic axis (OA) projection image has a circular arrangement of the scleral fibers around the ON (arrow). The circular arrangement exists just around the ON within the PPA. Outside the circular arrangement, the inner scleral fibers extend radially from the ON toward the fovea. The OA images (C, F, and I) are colored based on the color wheel in the lower right corner, with blue, yellow, pink, and green indicating the horizontal, vertical, and 2 oblique scanning directions, respectively. D: Fundus photograph of the left eye of a 51-55-year-old woman with a refractive error of -17.8 D and an AL of 31.4 mm showing diffuse choroidal atrophy with large PPA. E: *En face* OCT intensity projection image showing a large PPA. F: OA projection image shows a relatively small circular arrangement just around the ON (arrow) within the PPA. G: Fundus photograph of the right eye of a 51-55-year-old man with a refractive error of -10.6 D and an AL of 28.9 mm showing diffuse choroidal atrophy. H: *En face* OCT intensity image shows horizontally oval ON surrounded by annular PPA. I: OA projection image shows horizontally arranged oval fibers just around the ON (arrow) within the PPA.

### Circular arrangements of scleral fibers around optic nerve (ON)

In the *en face* OA images, the scleral fibers were noted to be arranged circularly around the ON in all eyes. The size and shape of this circular arrangement were determined solely by the ON, irrespective of the presence or size of the peripapillary atrophy (PPA; Figure 1). Even in eyes with a large PPA or deformed optic discs, the circular arrangement remained confined to the area immediately surrounding the ON. The circular arrangement appeared to be a projection of the outer sclera at the edge of the ON in the OA images (Figures 2–5) as reported in an earlier study.^12^ The fibers were composed essentially of the outer circumferentially oriented sclera fibers and were only seen and distinguishable in the OA images.

**Figure 2.**
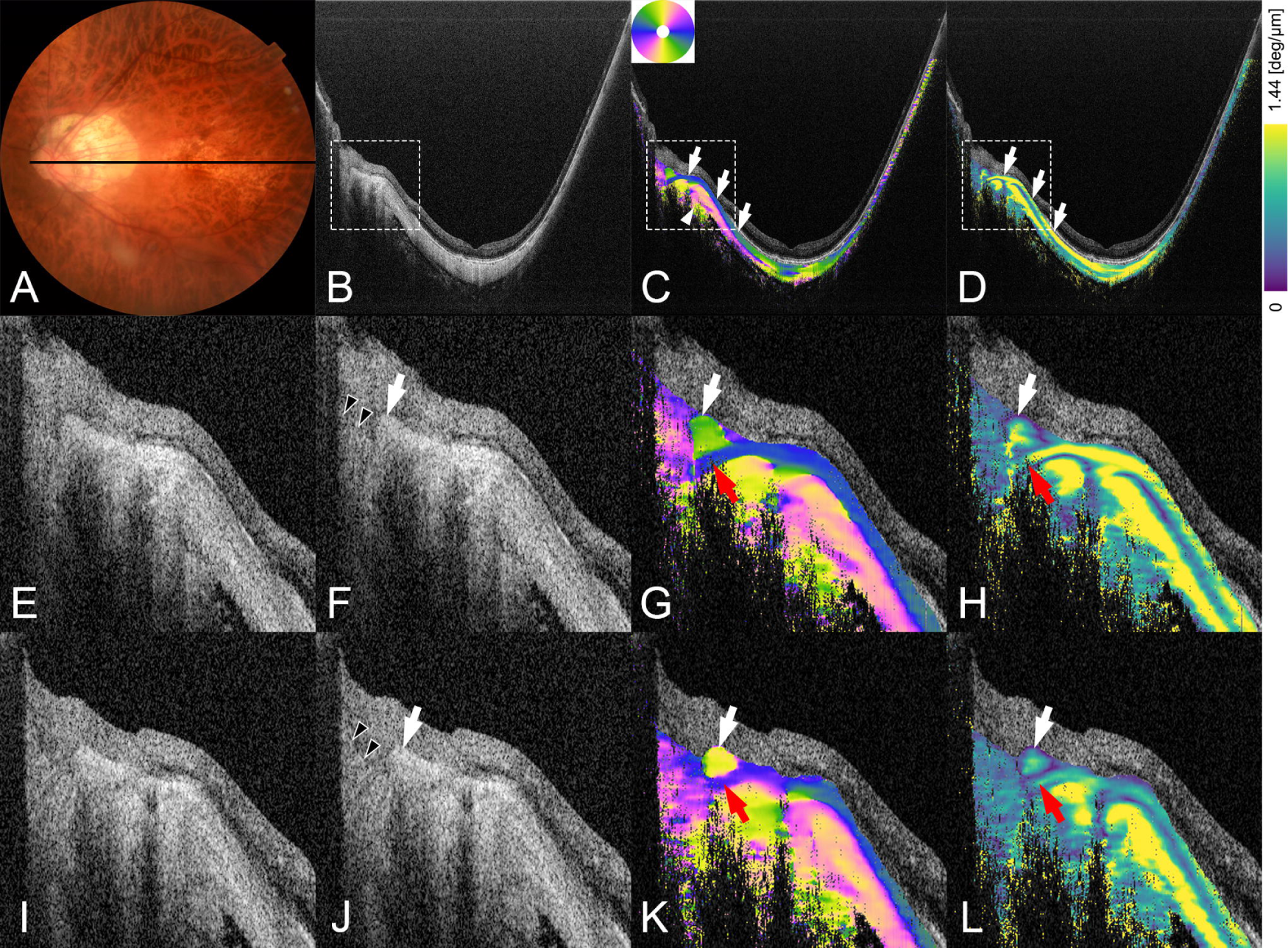
The circular arrangement of the scleral fibers around the optic nerve (ON) consists of the outer sclera with the inner sclera penetrating through this circle to continue toward the lamina cribrosa (LC). A: Fundus photograph of the left eye of a 61-65-year-old man with an implanted intraocular lens and an AL of 30.5 mm showing diffuse choroidal atrophy. The black line is the scanned line for the PS-OCT images in B to H. B:, C:, and D: Horizontal scan PS-OCT images across the fovea. B: Intensity image. C: OA image shows that the inner and outer layers of the sclera around the ON have different orientations of the scleral fibers. The inner sclera is composed mainly of radial fibers which are horizontally oriented in this cross-sectional image (blue; arrows). In contrast, the outer sclera is composed of circumferentially oriented fibers and is obliquely oriented in this section (mixed colors; arrowhead). D: Birefringence image showing that the inner scleral fibers have high birefringence (arrows). Peripapillary structures outlined by the squares are magnified in images E to H. E: to H: Magnified images of the squares in images B to D. E: and F: Intensity images show the anterior surface of LC (arrowheads). Peripapillary sclera appears to protrude anteriorly (arrow). G: In the OA image, the circular arrangement is located at the edge of the ON and is seen as the projection of the outer scleral fibers (white arrow). A penetration of almost the entire thickness of the inner sclera is observed (red arrow) beneath the circular arrangement and runs toward the LC. H: In the birefringence image, the circular arrangement shows high birefringence (white arrow), while penetrating inner scleral fibers have low birefringence (red arrow). I: to L: Peripapillary magnified PS-OCT images scanned at 235 μm inferior to the scan line shown in A. I: and J: Intensity images show the anterior surface of the LC (arrowheads). Peripapillary sclera appear to protrude anteriorly (arrow). K: In the OA image, the circular arrangement is located at the edge of the ON and is seen as a projection of the outer sclera (white arrow). A penetration of almost the entire thickness of the inner sclera is observed (red arrow) beneath the circular arrangement and runs toward the LC. L: In the birefringence image, the penetrating inner scleral fibers have low birefringence (red arrow). The OA images (C, G, and K) are colored based on the color wheel in image C, with blue, yellow, pink, and green indicating the horizontal, vertical, and 2 oblique scanning directions, respectively. The birefringence images (D, H, and L) are colored using the color bar in the upper right corner and are intentionally saturated to show the low birefringence structure more clearly.

### Penetration of inner sclera fibers within and below the circular arrangement

We found in the OA image that the inner scleral fibers did not terminate outside the circular arrangement. In 98 of 110 eyes (89.1%), the inner scleral fibers penetrated beneath the circular arrangement (Figure 2). The penetrating fibers ran toward the ON and probably toward the LC. In the remaining eyes, the penetration of the inner scleral fibers could not be detected because of an extremely tilted ON in 5 eyes, thick choroid in 4 eyes, and loss of the typical circular arrangement in 3 eyes.

Additionally, the inner scleral layer was observed to split axially into a Y-shape in 77 of 110 eyes (70.0%) at the initial point where the fibers penetrated the circular arrangement. However, in these eyes, the complete course of penetrating fibers was not consistently detected.

In representative cases (Figure 3), the superior branch passed over the body of the circular arrangement and extended toward the anterior portion of the LC. On the other hand, the inferior branch passed beneath the circular arrangement and extended into or beyond the LC.

**Figure 3.**
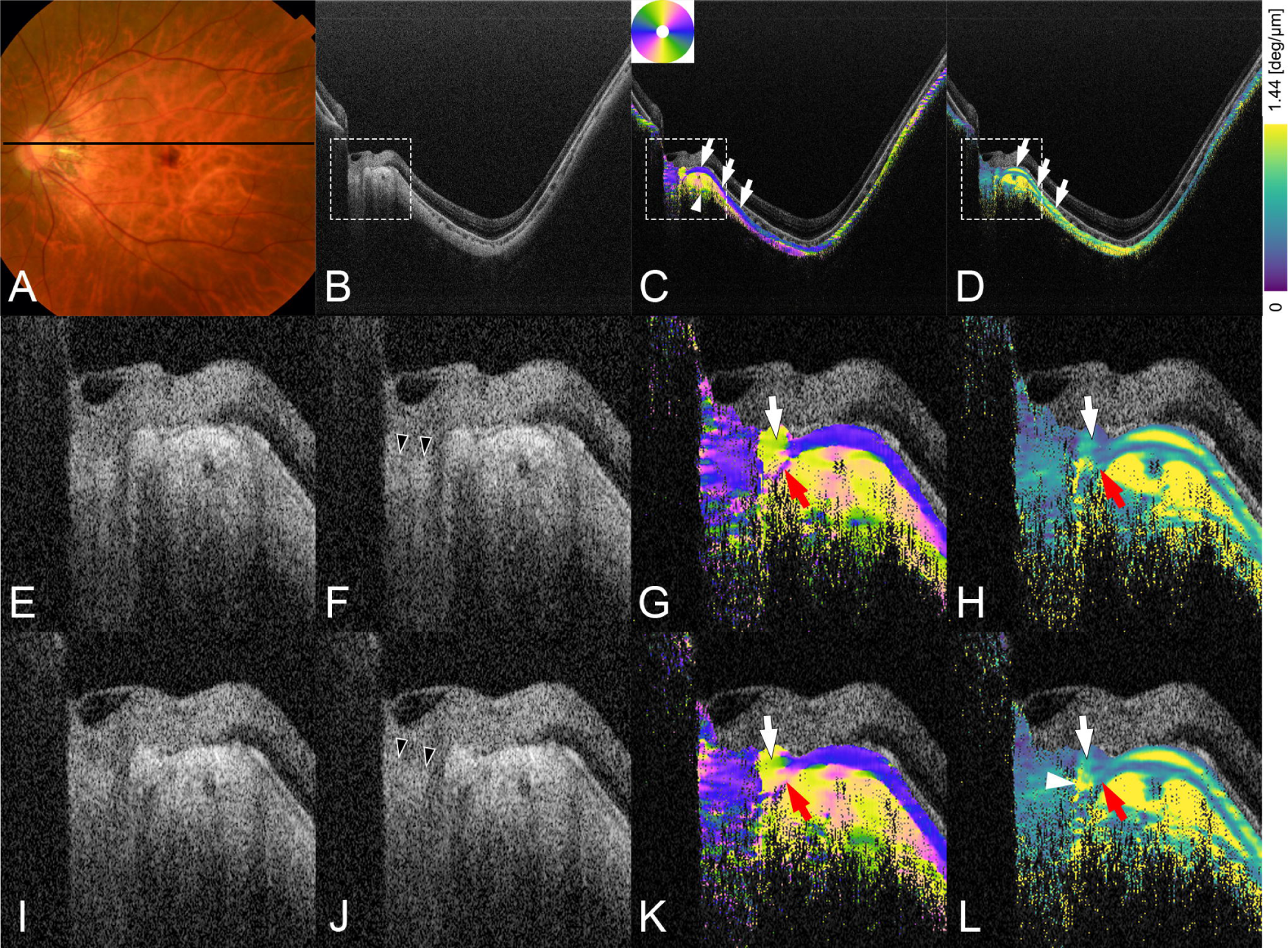
The circular arrangement of the scleral fibers around the optic nerve (ON) consists of the outer sclera. The inner sclera can be seen to penetrate through this circle to continue toward the lamina cribrosa (LC). The color wheel and bar labeling are the same as in Figure 2. A: Fundus photograph of the left eye of a 56-60-year-old woman with a refractive error of -19.0 D and an AL of 31.6 mm showing diffuse choroidal atrophy. Black line is the scanned line of PS-OCT images in B to H. B:, C:, and D: PS-OCT images correspond to the scan line shown in A. B: Intensity image. C: OA image showing that the inner sclera is horizontally oriented in this cross-sectional image (blue; arrows). The outer sclera is generally vertical in this section (yellow; arrowhead). D: Birefringence image shows that the inner scleral fibers are highly birefringent (arrows). Peripapillary structures in the squares are magnified and shown in images E to H. E: to H: Magnified images of the squares in images B to D. E: and F: Intensity images show the anterior surface of the LC (arrowheads). G: In the OA image, the inner scleral fibers do not terminate outside the circular arrangement; they split in a Y-shape pattern and penetrate and extend toward the LC. More specifically, the superior branch (white arrow) passes through the body of the circular arrangement and runs toward the anterior part of the LC. The inferior branch (red arrow) passes beneath the circular arrangement and extends into or deeper into the LC. H: Birefringence image showing that both the penetrating inner scleral fiber bundles have low birefringence (arrows). I: to L: Peripapillary magnified images of PS-OCT images scanned at 47 μm inferior to the scan line shown in A. I: and J: Intensity images show the anterior surface of the LC (arrowheads). K: In the OA image, the superior branch (white arrow) of the split inner scleral fibers penetrates within the body of circular arrangement and runs toward the anterior part of the LC. The inferior branch (red arrow) passes beneath the circular arrangement and extends into or deeper into the LC. L: Birefringence image. The circular arrangement shows high birefringence (arrowhead) but both of the penetrating inner scleral fiber bundles have low birefringence (arrows).

The inner scleral layer was also found to split into three or more branches. It was not divided into only two fiber bundles but it could be further subdivided into thinner bundles. Notably, 73 of 110 eyes (66.4%) had at least one small superior penetrating branch with its complete course extending through the main body of the circular arrangement. Because each eye had 256 PS-OCT image sections, some values overlapped, allowing a simultaneous detection of the above phenomena in a single eye.

In the birefringence images, all of the penetrating inner scleral fiber bundles had low birefringence.

### Resilience of circular and penetrating scleral fiber complexes

We also observed that even in cases of severe deformation of the peripapillary sclera, the circular arrangements and the penetrating inner scleral fibers remained unaffected (Figure 4). Various peripapillary abnormalities including conus pits and scleral ridges were commonly observed in eyes with pathologic myopia. Conus pits are thought to originate from scleral stretch-associated schisis or openings for the short posterior ciliary arteries that form pit-like clefts in the adjacent scleral crescent.^27,28^

**Figure 4.**
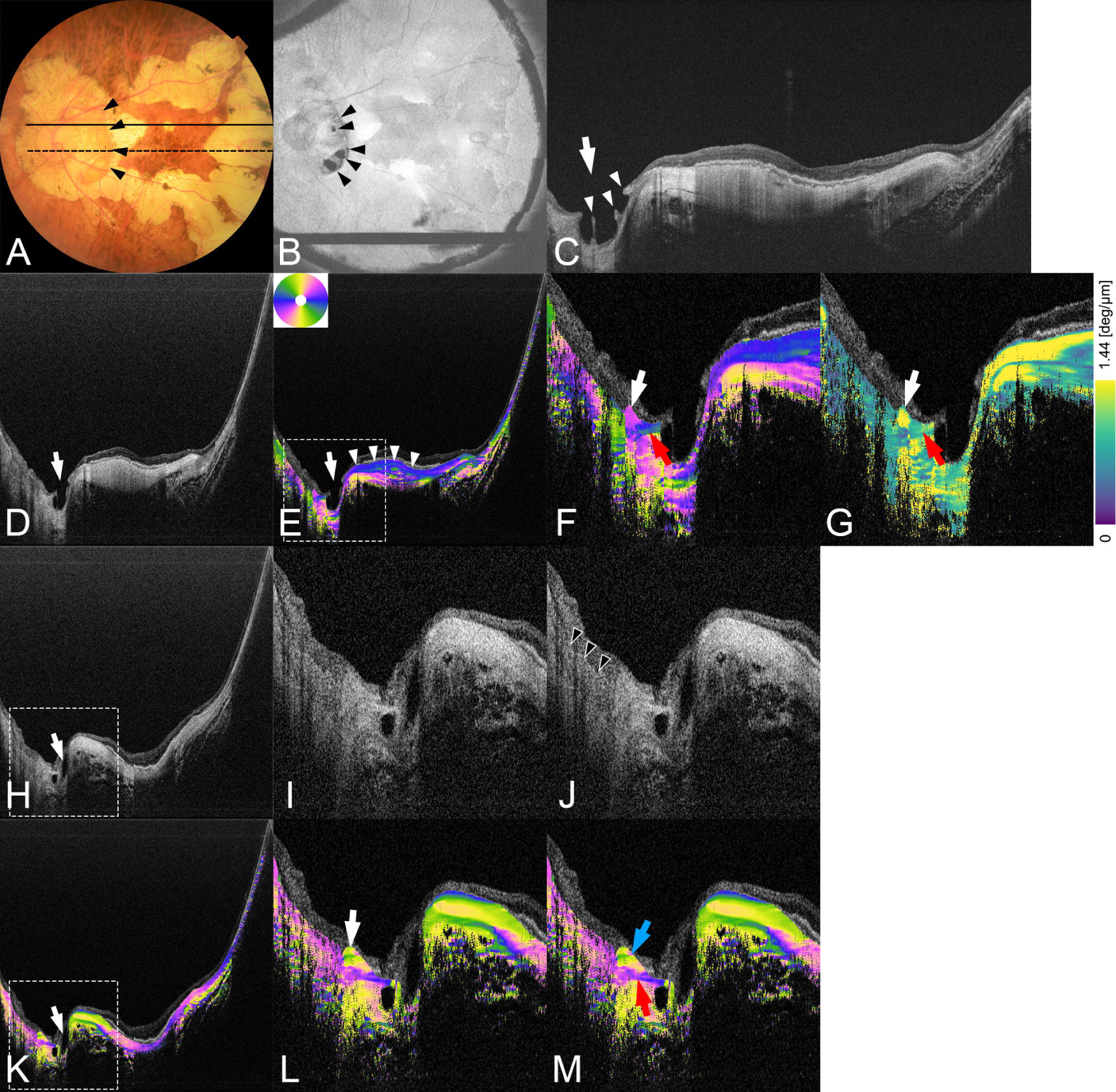
The inner scleral penetrating fibers remain even in eyes with a scleral conus pit. The color wheel and bar labeling are the same as in Figure 2. A: Left fundus of a 71-75-year-old woman with an implanted intraocular lens and an AL of 34.9 mm. Multiple areas of patchy choroidal atrophy can be seen surrounding the macula. A large area of peripapillary atrophy (PPA) can be seen around the optic nerve (ON). The scleral ridge can be seen temporal to the ON (arrowheads). The solid and dotted lines show the locations of two PS-OCT scans. B: *En face* OCT intensity image. Multiple conus pits can be clearly seen (arrowheads). C: A horizontal scan of swept-source OCT image across the macula shows a large and deep conus pit (arrow). The sclera at the conus pit is posteriorly bowed while the remaining retinal vessels remain protruded (arrowheads). Macular sclera shows a dome-shaped macula (DSM) appearance. D: to G: PS-OCT images corresponding to the dotted scan line shown in A. D: The intensity image shows a deep conus pit (arrow). DSM is also seen. E: OA images. The inner scleral layer (arrowheads) is thick at the DSM and continues until the conus pit (arrow). F: This is a magnified image of the square area in E. A projection of the outer scleral oblique fibers is seen just around the ON (white arrow). Penetration of horizontally running fibers just below the circular arrangement (red arrow) is seen toward the ON. G: Birefringence image shows high birefringence at the circular arrangement around the ON (white arrow) while penetrating inner scleral fibers show low birefringence (red arrow). H: to M: PS-OCT images corresponding to the solid scan line shown in A. H:, I:, and J: Intensity images. A conus pit is seen (arrow in H). I and J are the same magnified images of square area in H. The anterior surface of the LC is shown (arrowheads in J). K: OA images. A conus pit is seen (arrow in K). L and M are the same magnified images of the square area in K. L: Inner scleral layer is thin in this scan and ends before the conus pit. However, just around the ON, the projection of outer sclera (consisting of circular arranged fibers) is seen (arrow). M: The inner scleral fibers that penetrate the circular arrangement remain in place (two branches; blue and red arrows). The superior branch (blue arrow) passes through the body of the circular arrangement and runs toward the anterior part of the LC, and the inferior branch (red arrow) passes beneath the circular arrangement and extends into or beneath the LC.

**Figure 5.**
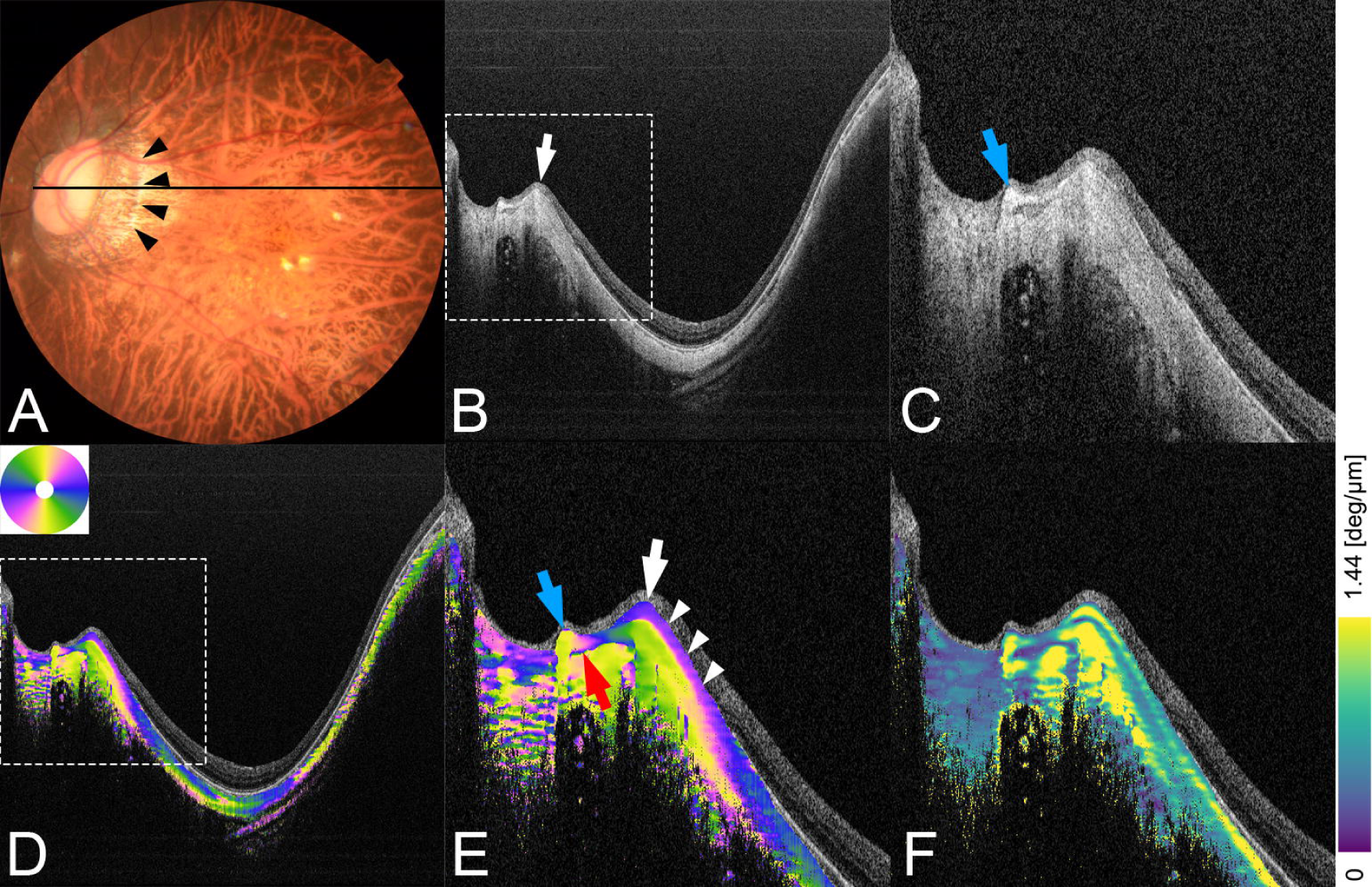
PS-OCT images of an eye with a scleral ridge. The color wheel and bar labeling are the same as in Figure 2. A: Left fundus of a 36-40-year-old woman with a refractive error of -17.4 D and an AL of 30.8 mm showing diffuse choroidal atrophy. Peripapillary atrophy is seen temporal to the optic nerve (ON). A scleral ridge can be seen temporal to the ON (arrowheads). The black line is the scanned line of the PS-OCT images in B to F. B: and C: In the intensity images of PS-OCT, the scleral ridge is clearly seen (white arrow). A magnified image of the square is shown in C. A slight protrusion of the sclera is also seen just around the ON (blue arrow). D: and E: In the OA images, the scleral ridge is seen as a protrusion of the entire scleral thickness rather than just the inner sclera. The scleral ridge is composed primarily of the outer rather than the inner scleral fibers. In the magnified image E, the scleral ridge is located outside the circular arrangement (white arrow). The inner sclera over the ridge is significantly thinned (arrowheads). There is another protrusion just around the ON which is the circular arrangement of the scleral fibers (blue arrow). The inner scleral fibers that penetrate posterior to this circle can also be seen (red arrow). F: Birefringence image of the same area. Scleral ridge and the circular arrangement have generally high birefringence.

Scleral ridges are anterior protrusions of the sclera located temporal to the optic disc.^23,28^ Among the 110 eyes, nine had conus pits with a circular arrangement around the ON in all of these eyes. In eight of them, the penetration of the inner scleral fibers remained in place, and one eye could not be examined in detail because of the loss of the typical circular arrangement.

### PS-OCT images of eyes with scleral ridge

Scleral ridges were observed in 27 of 110 (24.5%) eyes. In all 27 eyes, the protrusions involved the entire scleral thickness rather than only the inner sclera (Figure 5). In addition, the ridges were composed mainly of outer scleral fibers rather than the inner scleral fibers. The ridges were always located outside the circular arrangement of the scleral fibers.

## Discussion

The OA in the posterior sclera have been studied in rats, guinea pigs, and humans.^10–12,25^ These images consistently revealed a two-layer structure in the peripapillary sclera: an inner layer of radial fibers and an outer layer of circumferential fibers. The OA images from PS-OCT were needed to show the scleral fiber alignment. This imaging approach showed a clear difference between the inner and outer scleral layers (Figure S1) which then provided valuable information on the physiological and pathological states of the sclera *in vivo*. Building on the results of our laboratory findings on the characteristics of dome-shaped maculas (DSMs),^10^ we examined the fiber structure surrounding the ON in this study.

Our images showed a circular arrangement of the collagen fibers around the ON in all eyes, and they appeared as projections of the outer sclera at the edge of the ON (Figures 2-5). The circumferential arrangement of the fibers around the ON has been documented earlier by *in vitro* using techniques such as WAXS^5^, SALS^6^, PLM^7,13^, and more recently by *in vivo* PS-OCT images.^11,12^ The circumferential fibers adjacent to the ON form a proximal ring which we refer to as a circular arrangement. Our observations closely agree with previous findings, and the circular arrangement appeared more distinct and easier to identify in our cross-sectional OA images compared to those obtained by earlier algorithms.^7,12,13^

The schematic illustration in Figure S2 is based on our observations and shows the course of the peripapillary scleral fibers and the penetration pattern of the inner radial fibers. Previous histological studies reported that the outer sclera transitions into the dura mater while the inner sclera forms the LC and contributes to the pia mater.^29,30^ On the other hand, the earlier studies reported that the radial inner fibers emerged outside the circular arrangement slightly farther from the optic canal.^5–7,11–14^ However, how the inner sclera is connected to the LC has not been conclusively determined.

In 89.1% of the eyes studied, cross-sectional OA images showed that the inner radial scleral fibers penetrated beneath the circular arrangement and ran toward the ON and the LC (Figure 2). A Y-shaped split was seen in 70.0% of eyes at the initial point of penetration. In addition, the radial fibers were seen to divide into multiple bundles, and 66.4% of the eyes had at least one fully visible superior bundle (Figures 3 and 4).

Our findings are the first detailing the unique patterns of inner scleral fibers that penetrated the ring and extended toward the ON rather than terminating outside. These findings provide new information on the contribution of the inner sclera to the formation of the LC. All penetrating bundles had low birefringence (Figures 2H, 2L, 3H, 3L, 4G), indicating less aligned and more interwoven scleral fibers.

The structural and functional relationships between the circumferential and radial fibers have not been determined. Ji et al. reported discrete LC insertions into the sclera with significant variations in the depth and quadrant distribution.^31^ Our findings showed that the inner scleral fibers penetrated the circular arrangement and split into superior and inferior or multiple branches, and closely aligned with these depth-dependent variations in the LC insertions.

In addition, the inconsistent observations of this penetration across cross-sections reflect the discrete nature of the LC insertions. These spatial correlations strongly support our suggestion that the radial fibers of the inner sclera directly connect with the LC by this penetration mechanism. Most previous studies focused on *en face* observations, with few employing continuous cross-sectional approaches by PS-OCT. Our study has bridged this methodological gap, leading to novel findings.

The circumferential fibers are widely accepted as protection for neural tissues by resisting canal expansion and providing structural support for the ON.^5,7,13,14^ Hua et al.^1^ showed that the radial fibers reduced the displacement of the LC and the peripapillary scleral tension under elevated IOP through finite element models. The combination of radial and circumferential fibers protected the ON better than either alone.

The *en face* OA images showed that the size and shape of the circular arrangement depended exclusively on the ON, which was unaffected by the presence or size of PPA (Figure 1). Cross-sectional OA images showed that even significant deformations of the peripapillary sclera, such as a scleral pit, did not alter the integrity of the complexes of the circular arrangements and the penetrating inner scleral fibers (Figure 4). These findings suggested that the structural stability of the circular arrangement, and emphasized the critical support resulting from its interweaving with the penetrating fibers. The penetrating fibers may act as anchors, stabilizing the circular arrangement through a fabric-like interweaving which would provide mechanical support.

Scleral ridges were observed in 24.5% of eyes as full-thickness protrusions and were not limited to the inner sclera. They consisted primarily of outer scleral fibers (Figure 5). Previous research from our laboratory using PS-OCT showed that the inner scleral fibers aggregated without a thickening of the outer sclera in highly myopic eyes with DSM sites.^10^ Although both scleral ridges and DSMs involved an inward protrusion of the sclera, their structural components were markedly different. The distinct compositions of the scleral abnormalities suggested different pathogenic mechanisms potentially requiring individualized management for each type of scleral abnormality.

Our study has several limitations. First, it included only myopic eyes and mostly highly myopic eyes as full-thickness scleral imaging was feasible only with significant choroidal thinning. Because scleral fiber penetrations were observed in most cases, we propose that the complexes likely represented a physiological structure in all eyes. Future basic studies are needed to confirm these findings. Second, the LC region was not suitable for study because of the spatially unresolved birefringent signals from the retinal nerve fibers.

Consequently, a direct confirmation that the penetrating fibers form the LC was not achieved, but SS-OCT indirectly validated this relationship by locating the anterior surface of the LC. Third, the imaging principles of OA, relying on the preferential orientation of net birefringence, could not fully represent the complex microstructure of the sclera, such as the individual collagen lamellae^22^ and the waviness of the collagen fibers^14^. Fourth, consistent with Willemse et al.^12^, we also failed to detect regions of interweaving fibers throughout the scleral depth described by Gogola et al.^7^ Last, the pia mater that is situated parallel to the incident light, could not be viewed through its OA, blocking an exploration of its relationship^29,30^ with the inner radial scleral fibers. SS-OCT has difficulties in identifying the posterior LC surface, although some of the penetrating fibers appeared to extend posteriorly to the LC (Figure S2). These fibers possibly linked the inner scleral fibers to the formation of the pia mater. However, current technology cannot verify this. These technical limitations underscore the need for future advancements in the algorithms to optimize PS-OCT which would then enable a broader application in clinical practice.

In conclusion, we examined the structural characteristics of the peripapillary sclera by PS-OCT, and we found that the circular arrangement of the scleral fibers around the ON is stable. The inner radial fibers penetrate and extend toward the LC and act as anchors of the circular arrangement. Scleral ridges had distinct compositional differences compared to DSMs, suggesting that the mechanisms underlying peripapillary scleral abnormalities differ from those in other regions. These findings offer insights for future clinical management.

## Data Availability

All data produced in the present study are available upon reasonable request to the authors.

