## Supplementary figures for "Penetration of Inner Scleral Fibers into Peripapillary Sclera Revealed by Polarization-Sensitive OCT"

**Figure S1**. An example of scleral fiber arrangement in a highly myopic eye with mild diffuse atrophy.

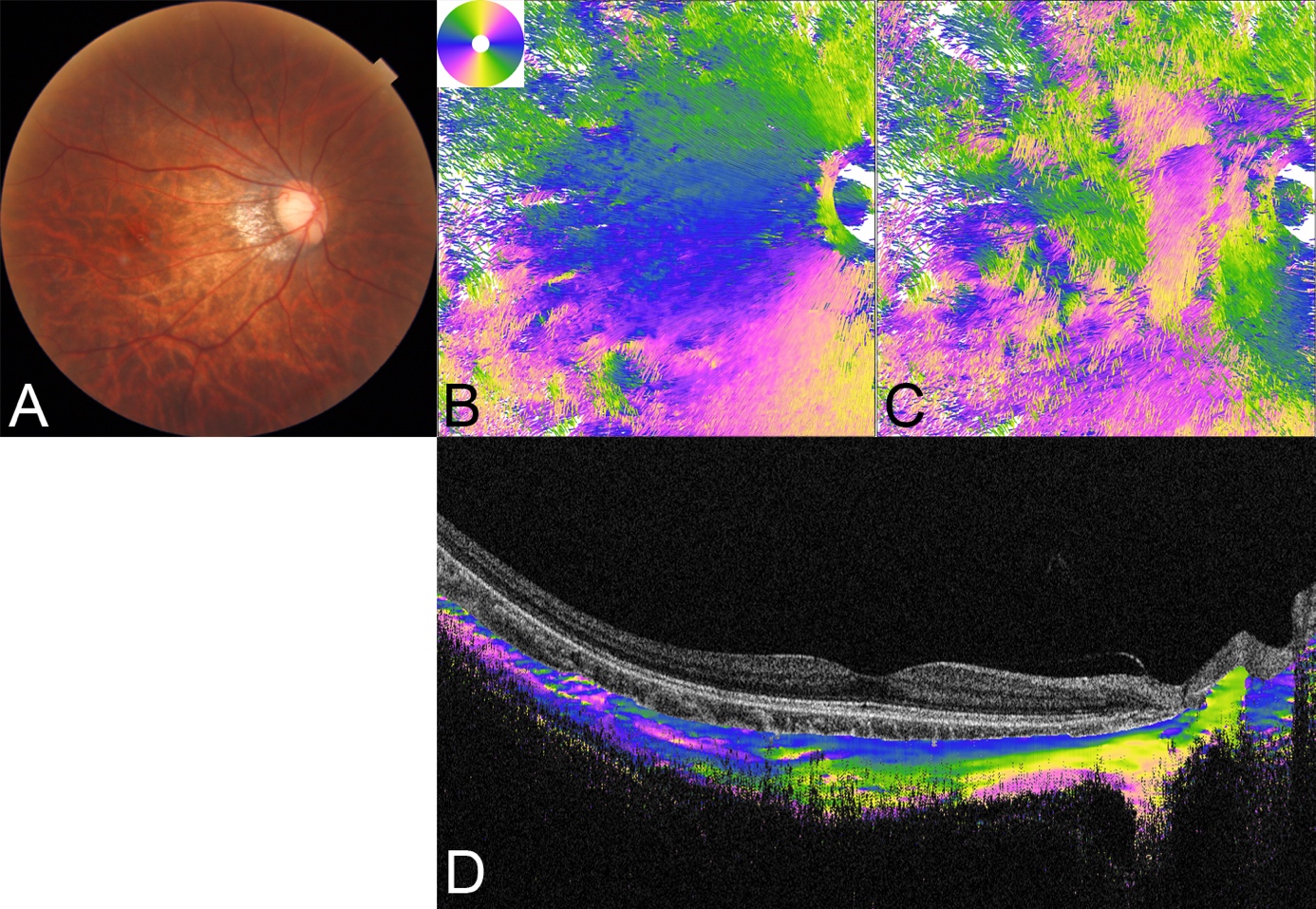

A: Fundus photograph of the right eye of a 21-25-year-old woman with a refractive error of -13.5 diopters (D) and an axial length (AL) of 30.7 mm showing diffuse choroidal atrophy. The color wheel labeling is the same as in Figure 2.

B: and C: Streamlined images from both the interior and exterior of the eye, rendered using ParaView. The images clearly show the course of the scleral fibers.

B: The peripapillary region shows a circular arrangement of fibers surrounding the optic nerve (ON), with inner scleral fibers extending radially from this circle toward the periphery.

C: The outer scleral fibers have vertical and oblique orientations.

D: The optic axis (OA) image across the fovea shows that the inner scleral fibers are arranged horizontally, coded in blue, while the outer scleral fibers are mainly arranged vertically, coded in yellow.

**Figure S2.** Schematic illustration of the course of the peripapillary sclera.

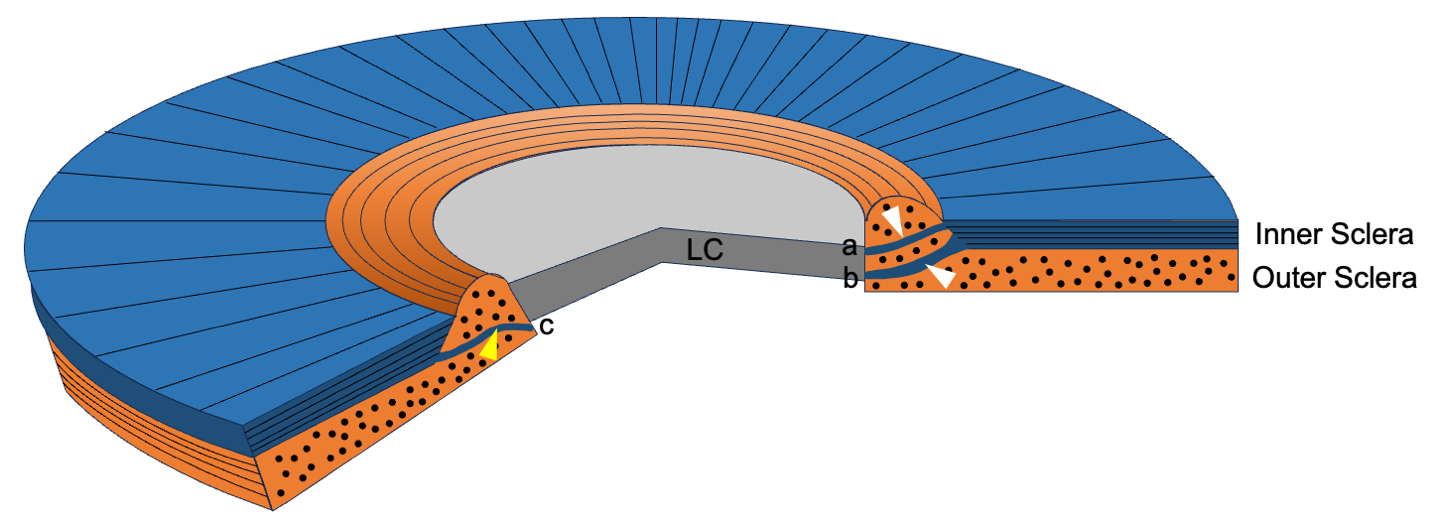

The inner and outer layers of the sclera around the ON have different orientations. The inner scleral fibers are radially oriented (blue layer) while the outer scleral fibers are circumferentially oriented (orange layer). Because the inner radial fibers are located slightly farther from the ON, the outer concentric fibers can be observed as a ring-like arrangement (circular) just around the edge of the ON in the *en face* OCT OA projection image. This circular arrangement is a protrusion of the outer sclera. The inner scleral fibers do not terminate outside the circular arrangement; some fibers split and penetrate into the superficial portion and deeper portion of the LC (white arrowheads; fibers a and b in the right section). Some penetrating fibers extend posteriorly to the LC, which may explain how the inner scleral fibers form the pia mater (yellow arrowhead; fibers c in the left section).
